# Cancer as a novel Risk Factor for Major Adverse Cardiovascular Events in Secondary Prevention

**DOI:** 10.1101/2024.07.16.24310536

**Authors:** Renzo Melchiori, Sara Diaz Saravia, Pablo Rubio, Lucas Szlaien, Romina Mouriño, Martin O’Flaherty, Miguel Rizzo, Alejandro Hita

**Affiliations:** Department of Cardiology and Echocardiography, Hospital Universitario Austral, Austral University, Buenos Aires, Argentina; Department of Medicine, Icahn School of Medicine at Mount Sinai; Department of Public Health, Policy and Systems, University of Liverpool, United Kingdom; Department of Hematology and Oncology, Hospital Universitario Austral, Austral University, Buenos Aires, Argentina

**Author notes:** **Corresponding Author:** Renzo Melchiori MD^1^, Department of Cardiology and Echocardiography, Hospital Universitario Austral, Austral University, Buenos Aires, Argentina.

## Abstract

**Introduction:** The inflammatory mechanisms of cancer can be associated with atherosclerosis development and progression. Although there is an incidence rate of cardiovascular events in secondary prevention following a first acute coronary syndrome (ACS).

**Methods:** A retrospective study cohort study including patients who underwent a PCI for first Acute Coronary Syndrome (ACS), and without prior history of Major Cardiovascular Events (MACE) from 2008 to 2022 was analyzed. Included patients were grouped according to the absence or presence of cancer: G1 non-oncologic, and G2 oncologic (either prior or actual history). We compared the incidence rate ratio of MACE within 3 years after STEMI between groups Time-to-event analysis was conducted through proportional Cox regression analysis, estimating hazard ratio, and corresponding 95% confidence intervals (95% CI)

**Results:** Out of 937 patients who underwent a PCI, 787 patients were included of which 88.7% (n=698) presented without cancer. Over a median follow-up time of 45 months [IQR= 14-72], the incidence rate of MACE was 4.4 cases per 1000 patients/months of follow-up (n=173 MACE events). When comparing both groups, the incidence rate ratio of MACE was 1.9 (95% CI 1.24-2.99), significantly increased in the cancer group (P=0.0032) without showing differences in median follow-up times. Cancer was an independent predictor of MACE HR 1.84 (95% CI 1.19-2.85; P=0.006), adjusted for hypertension, dyslipidemia, diabetes, smoking history, sedentary lifestyle, obesity, age, male gender, and family history of cardiovascular disease.

**Conclusions:** Patients with cancer represent a novel independent risk factor for MACE, even following secondary preventive therapies. These results highlight future endpoints for cardiovascular prevention and further public health interventions on this population.

## INTRODUCTION

Cancer is the second cause of death worldwide, only surpassed by cardiovascular diseases (1). In recent decades, cancer mortality rates have decreased, with a consequent increase in overall survival (2). For those patients who survive cancer, oncological treatments and cardiovascular and cerebrovascular diseases become the main causes of mortality (3, 4). In 2022, there were approximately 4.2 million cases of cancer in the region of the Americas, and this number is projected to increase up to 6.7 million in 2045 according to the Pan-American Health Organization. This growing number calls for a better understanding of the relationship of cancer and cardiovascular disease in the long term.

Multiple studies have emphasized the relationship between the pathophysiological mechanisms of cancer and cardiovascular disease (5-6). There is emerging evidence on the role of oncological disease as a precipitator of chronic inflammation and long-term endothelial damage (7-11). Although there are multiple registry publications in relation to the increase in cardiovascular events in the cancer population (3, 5, 17-21), the evidence for secondary prevention is scarce.

However, less is known in terms of the excess risk of cancer on Major Adverse Cardiovascular Events (MACE) in the context of secondary prevention. If this association exists, its role in modifying management and prognostic strategies will need to be defined, implying a significant impact in public health and cardiovascular prevention. Therefore, we sought to determine the independent association of oncological disease and its effect on the incidence of MACE compared to patients without cancer in the secondary prevention setting.

## METHODS

### Setting and Study Design

A retrospective cohort study was conducted including adult patients (>17 years of age) who were admitted to the Cardiac Care Unit at the Austral University Hospital with Acute Coronary Syndrome (ACS) from January 1^st^, 2009, to March 31^st^, 2023. This registry was formed through the review of Electronic Medical Records of patients presenting with ACS during that period, and subjects were selected according to the eligibility criteria defined (see section “Eligibility Criteria”). Institutional Ethical Approval was granted by the Ethics Board and written informed consent was obtained from the patients before their admission to the institution (Institutional Evaluation Committee of the Universidad Austral, CIE No. P21-022).

### Eligibility Criteria

Patients eligible for this study included those who underwent Left Heart Catheterization (LHC) as part of the therapeutic diagnostic strategy during their first hospitalization due to ACS. Patients with prior history of Myocardial Infarction (MI), or Cerebrovascular disease (CVA), or either percutaneous Coronary Intervention (PCI) or Coronary Artery Bypass Graft Surgery (CABG) were excluded.

### Exposure variable of interest

The exposure group was defined as any active oncological history (under treatment) or referred, regardless of its duration, prior to entry into the cohort, irrespective of the type of treatment received. The cancer history necessary for our study was obtained from the patient’s electronic medical records. It was characterized by the type of neoplasm, defining hematologic neoplasia as leukemia, lymphomas, myeloproliferative syndromes, and multiple myeloma, and solid organ tumors as any tumor that is non-hematological, establishing cellular type where possible. The years since diagnosis were determined as the time from cancer diagnosis to admission to the coronary unit or cardiovascular study as part of the admission. Oncological stage was established according to the TNM classification system.

Regarding cancer therapies, it was assessed whether patients were under active treatment or not. Active chemotherapy was defined as having received chemotherapy or immunotherapy within 90 days prior to initial hospital presentation. Specific treatment types were categorized into chemotherapy, immunotherapy, hormone therapy, and radiotherapy. Radiation therapy history was defined as radiation treatment to any organ, establishing the cumulative dose, if possible, as the oncological treatment may have been administered at another institution. It was also determined whether anthracyclines were used in the oncological treatment. The time since completion of treatment was established in years at the time of admission for acute coronary syndrome (ACS). Skin cancer basal cell carcinoma or squamous cell carcinoma were excluded.

### Other exposure variables of interest and co-interventions

The known risk factors were collected by the attending physician upon admission for the ACS in the electronic medical records, including hypertension defined as systolic blood pressure greater than or equal to 140 mmHg and/or diastolic blood pressure greater than or equal to 90 mmHg or the use of antihypertensive medications at the time of hospitalization. Diabetes mellitus was defined as HbA1c levels greater than 6.5 mg/dL in the laboratory and/or documented history in the medical record and/or use of insulin. Smoking history was considered, including former and active smokers. Dyslipidemia was defined as values above normal for total cholesterol, LDL cholesterol, and/or triglycerides, as well as the use of lipid-lowering medications. Obesity was defined as a body mass index equal to or greater than 30. Sedentary lifestyle was defined as less than 2.5 hours of exercise per week, and family history included the presence of myocardial infarction or stroke in a first-degree relative before the age of 55 for men or 65 for women.

The clinical variables and laboratory tests to assess adherence to secondary prevention goals were rate of statins rate, dosing of statins (standardized to equivalent doses of atorvastatin), LDL value (mg/dl), use of dual antiplatelet therapy (DAPT), duration of DAPT and use of Aspirin. These parameters were measured in both oncology and non-oncology patients as part of their follow-up.

### Primary outcome assessment, definition, and statistical analysis

The primary endpoint was the incidence rate of new MACE over the follow-up period since hospital discharge after the primary ACS. MACE was defined as a composite of death by cardiovascular cause, non-fatal Myocardial Infarction (MI), non-fatal Cerebrovascular Accident (CVA), and repeat non-staged Percutaneous Coronary Intervention (PCI). Cardiovascular death was considered when the patient was admitted due to a MACE. This does not include patients admitted for non-cardiovascular events who subsequently developed STEMI or CVA during their hospitalization. The non-fatal myocardial infarction was considered using the current universal definition of myocardial infarction at the time of admission (14).

Stroke was defined as compatible clinical symptoms plus imaging findings of brain injury on computed tomography or magnetic resonance imaging (15). In our study, scheduled angioplasty was considered for all PCI procedures performed after one month from the primary event due to symptoms or ischemia on stress test.

The surveillance follow-up for MACE in each patient was conducted by the Primary Care Physician (PCP) or the Principal Cardiologist. Patients who were not followed up at our institution were contacted by phone by medical personnel who underwent a structured interview. (Table S5)

The secondary endpoint was the comparison of incidence rates of MACE in subpopulations of patients without prior radiotherapy in any body region, and in patients without active chemotherapy at the time of ACS. The objectives of secondary prevention were also derived from the Electronic Medical Record (EMR) data and were aligned with the current guidelines of the American College of Cardiology (ACC) and the European Society of Cardiology (ESC), conducted by the Primary Care Physician (PCP) or the Principal Cardiologist. These parameters were measured in both oncological and non-oncological patients as part of their follow-up.

We estimated a sample power for time-to-event analysis, considering an alpha and beta error of 0.05 and 0.2, respectively. Following the Friedman method, a total number of 167 MACE over the follow-up were necessary to obtain a power of 80%. Categorical variables are reported as a frequency and percentage and subgroups were compared using Chi-square or Fisher exact test, as appropriate. Continuous variables are reported as mean and standard deviation (±SD) or as median (IQR), and were compared using the t test or Wilcoxon, for normal or non-parametric distribution, respectively. To determine whether cancer acted as an independent predictor of MACE, adjusted for traditional cardiovascular risk factors, a Cox regression model was conducted. Hazard ratios and its corresponding 95%CI were estimated. Performance of the final model was evaluated through calibration (observed vs expected outcomes) and discrimination with Harrell’s c-statistics index. Kaplan-Meier survival curves were computed and compared using the log-rank test. The same analysis was performed with the secondary objective subpopulations. All statistical analyses were performed with Stata® 17.0BE.

## RESULTS

An initial screening process detected 937 patients who had presented to our hospital for ACS and had undergone a LHC as part of their work-up. Of those patients, we excluded 35 patients who had undergone CABG, 31 CVA, prior myocardial infarction 23, death 21, and 40 patients were lost to follow-up. These patients who had been lost to follow-up were looked-up on national registries to ensure they were not deceased. A total of 787 patients who completed follow-up were included, 698 (88.7%) without cancer (G1) and 89 (11.3%) with a history of cancer or active cancer (G2). The median follow-up time for the entire cohort was 48 months (IQR=15-72), being 48 months (IQR=15-84) for G1 and 36 months (IQR=11-48) (0.03).

Patients with cancer were significantly older (67 years/±10 vs 60.2 years ±11, p 0.0001) and had a higher prevalence of insulin-dependent diabetes mellitus (10% vs 4%, p=0.01). The rest of the baseline characteristics can be seen in Table 1. The therapeutic strategies and general therapeutic objectives for the management of ACS were adjusted to the guidelines available at that time (Table S1). There were no significant differences in LHC findings between the two groups, as seen on Table 1.

**Table 1.** Baseline Characteristics of both subpopulations at the time of ACS presentation.

| Variables | Total (N=787) | G1 (N=698) | G2 (N=89) | P Value |
| --- | --- | --- | --- | --- |
| Age (years) | 61 (SD $\pm 11$) | 60.2 (SD $\pm 11$) | 67 (SD $\pm 10$) | 0.0001 |
| Male Sex | 645 (82%) | 572 (81.9%) | 73 (82%) | 0.98 |
| Hypertension | 496 (63%) | 441 (63.2%) | 55 (61.8%) | 0.78 |
| No DM | 625 (79.4) | 563 (80.5%) | 63 (71%) | 0.03 |
| NIDDM | 125 (17.5%) | 108 (15.5%) | 17 (19%) | 0.38 |
| IDDM | 37 (4.7%) | 28 (4%) | 9 (10%) | 0.56 |
| History of Smoking | 480 (61%) | 430 (62%) | 50 (56%) | 0.32 |
| Dyslipidemia | 494 (62.7%) | 442 (63.2%) | 52 (58.4%) | 0.33 |
| Hyperuricemia | 19 (2.41%) | 16 (2.29%) | 3 (3.7%) | 0.53 |
| Obesity | 290 (36.9%) | 267 (38.3%) | 23 (25.8%) | 0.022 |
| Family History of Cardiovascular Disease | 137 (17.4%) | 130 (18.6%) | 7 (7.9%) | 0.012 |
| Sedentarism | 441 (56%) | 405 (58%) | 36 (40.4%) | 0.002 |
| Length of Stay (days) | 3 (IQR 2-5) | 3 (IQR 2-6) | 4 (IQR 3-5) | 0.1 |
| Mean LVEF at time of Discharge (%) | 54 ($\pm 10.5$) | 54 ($\pm 10$) | 53 ($\pm 11$) | 0.44 |
| **Number of Affected vessels** |  |  |  |  |
| 0 | 72 (9.15%) | 61 (8.74%) | 11 (12.3%) | 0.26 |
| 1 | 338 (42.9%) | 306 (43.8%) | 32 (35.9%) | 0.15 |
| 2 | 277 (35.2%) | 244 (34.9%) | 33 (37.1%) | 0.69 |
| 3 | 100 (12.7%) | 87 (12.5%) | 13 (14.6%) | 0.59 |
| Left Main | 20 (2.54%) | 18 (2.58%) | 2 (2.25%) | 0.851 |
| Left Anterior Descending | 475 (60.4%) | 421 (60.3%) | 54 (60.7%) | 0.94 |
| Left Circumflex | 314 (39.9%) | 276 (39.5%) | 38 (42.7%) | 0.56 |
| Right Coronary | 363 (46.1%) | 315 (45.1%) | 48 (53.9%) | 0.11 |
| **Number of Treated Lesions** |  |  |  |  |
| 1 | 405 (51.4%) | 368 (53.5%) | 37 (41.6%) | 0.05 |
| 2 | 239 (30.4%) | 209 (30.3%) | 30 (33.7%) | 0.58 |
| 3 | 70 (8.89%) | 59 (8.6%) | 11 (12.3%) | 0.224 |
| **BARC Score** |  |  |  |  |
| 0 | 579 (73.6%) | 476 (68.2%) | 67 (75.3%) | 0.74 |
| 1 | 188 (23.9%) | 202 (28.9%) | 20 (22.5%) | 0.69 |
| 2 | 16 (2.03%) | 15 (2.15%) | 1 (1.12%) | 0.51 |
| 3 | 4 (0.51%) | 3 (0.43%) | 1 (1.12%) | 0.32 |

G1= Non-Oncologic Group, G2= Oncologic Group, DM= Diabetes Mellitus, NIDDM= Non-Insulin Dependent Diabetes Mellitus, IDDM= Insulin-Dependent Diabetes Mellitus.

The median time from cancer diagnosis to cohort entry was 24 months for G2. The cell line and staging characteristics of the oncology population can be seen in Table S1-S2. The objectives and management of secondary prevention were aligned with the guidelines in effect at the time of MACE. There were no statistically significant differences between LDL values, statin adherence, statin dose, DAPT and aspirin use between the two groups at the time of MACE (Table S4).

### Incidence

A total of 173 (21.9%) MACE were identified in the follow-up period in both groups combined. The cumulative incidence of MACE for G1 was 21.06% (147/698) and for G2 was 29.3% (26/89). G2 presented a significantly higher incidence density of MACE when compared to G1 [8 MACE/1000 patients/month (95% CI 5.5-11.8) vs 4 MACE/1000 patients/month (95% CI 3. 4-4.8), p=0.0032]. The rate of cardiovascular death during the follow-up period was 4.5% for G2 and 0.14% for G1 (P=0.001). Other findings on the MACE events during the follow-up period can be found on Table 2. The non-cardiovascular mortality of the entire cohort during the follow-up period was 1.01%. In G2, non-cardiovascular death was higher than in G1 [5 (5.62%) vs 3 (0.43%), p = 0.001].

**Table 2.**
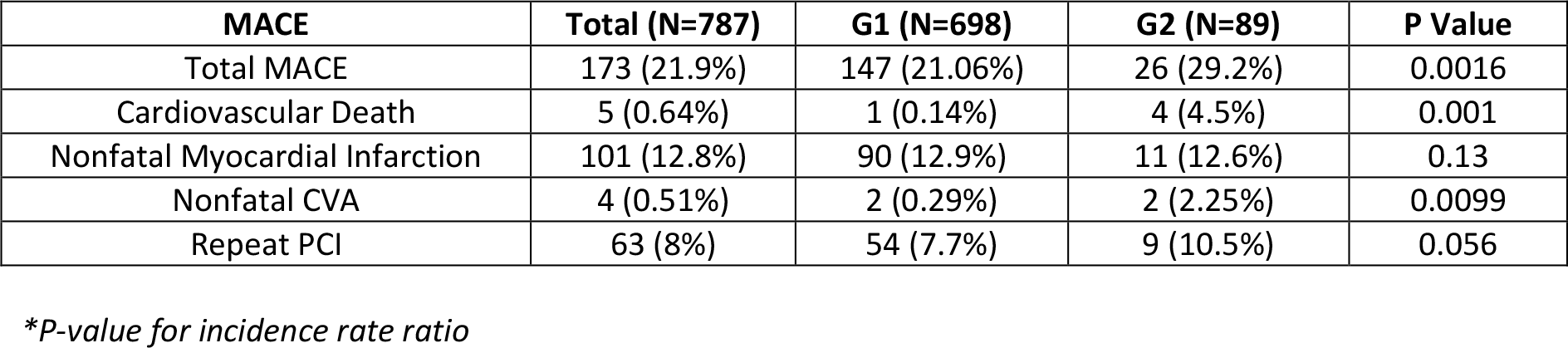
Incidence of Major Adverse Cardiovascular Events.

**Table 3.** Multivariate Analysis for MACE.

| Predictors for MACE | Hazard Ratio | 95% Confidence Interval | P Value |
| --- | --- | --- | --- |
| History of Cancer | 1.84 | 1.2-2.84 | 0.006 |
| Smoking history | 1.21 | 0.86-1.69 | 0.25 |
| Diabetes Mellitus | 1.29 | 0.90-1.86 | 0.16 |
| Age | 1.01 | 0.99-1.02 | 0.23 |
| Male Sex | 1.36 | 0.87-2.12 | 0.17 |
| Dyslipidemia | 1.1 | 0.79-1.54 | 0.56 |
| Obesity | 0.88 | 0.63-1.23 | 0.46 |
| Hypertension | 1.68 | 1.17-2.40 | 0.005 |
| Sedentarism | 1.13 | 0.80-1.60 | 0.46 |
| Family History | 0.75 | 0.48-1.18 | 0.22 |

The Kaplan Meier curve showed a statistically significant difference in time-to-MACE between the two groups, with p=0.0015 (Figure 2) and a LR Chi 32.7.

**Figure 1.**
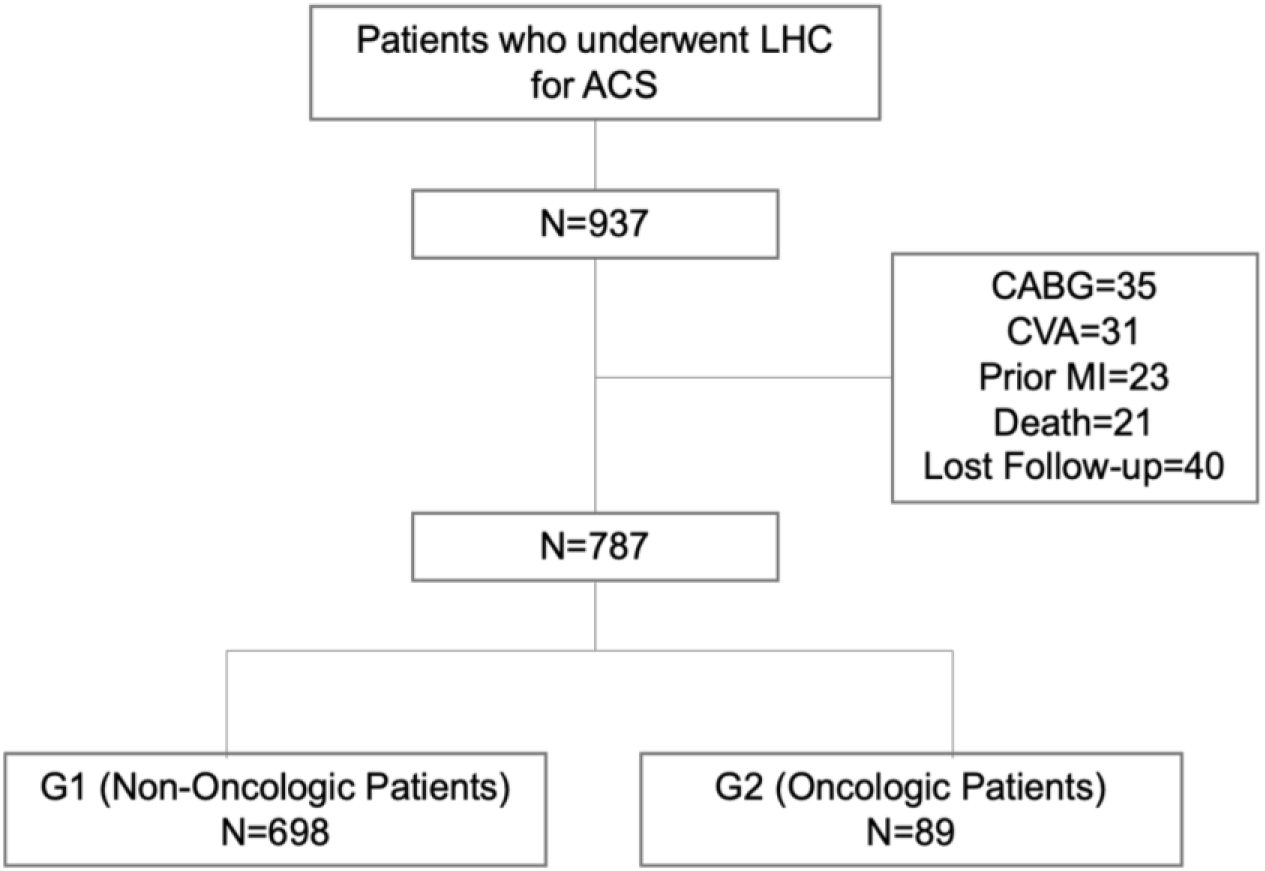
Flow chart of patient selection

**Figure 2.**
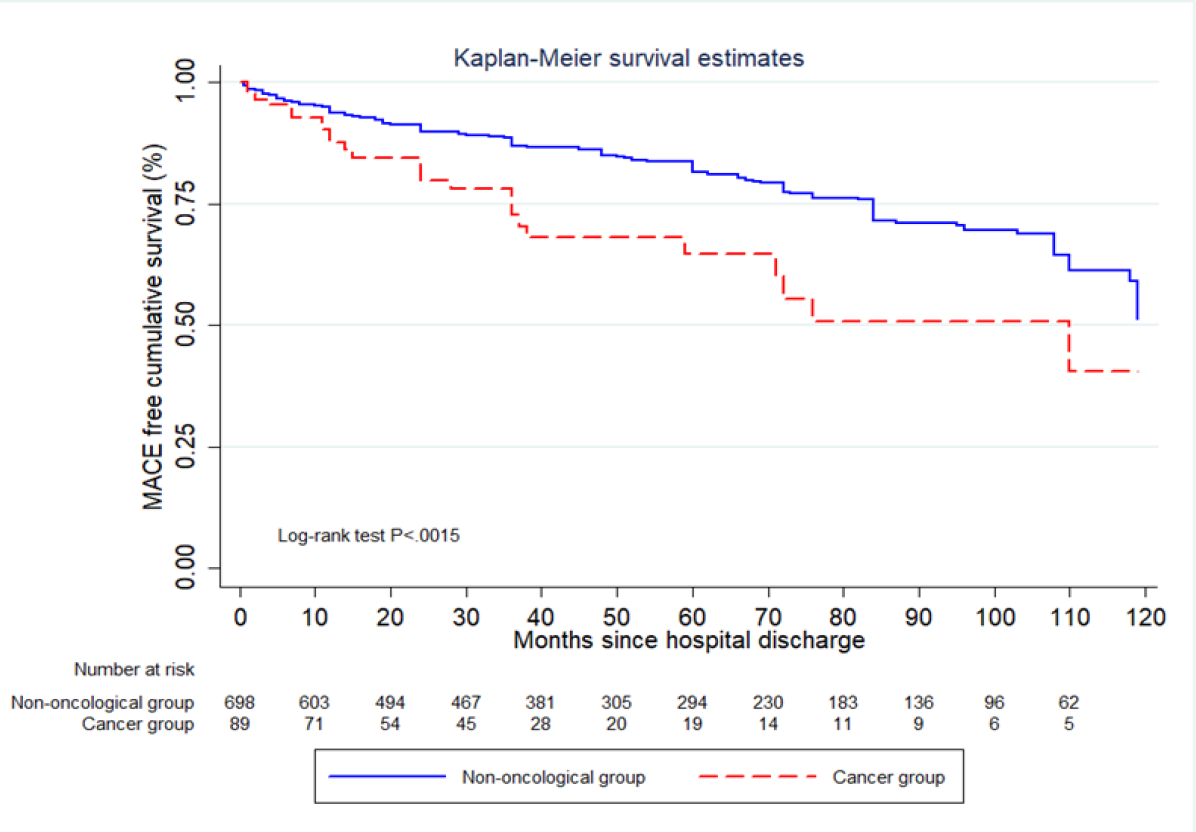
Kaplan Meier Curve for Time-to-MACE.

Multivariate Cox hazard analysis showed that cancer was an independent predictor of MACE (HR 1.84, [95% CI 1.19-2.85] p=0.006, Harrell’s C p=0.63), adjusted for hypertension, dyslipidemia, diabetes, history smoking, sedentary lifestyle, obesity, age, sex, and family history of cardiovascular disease.

### Secondary Outcomes

A total of 65 patients with a history of cancer without prior radiotherapy were included in the subgroup analysis. The Cox analysis showed that Hazard Ratio (HR) of 1.92, (95% CI 1.19-3.08) p=0.007, adjusted for age, sex, and the classic risk factors for the oncology population. The Kaplan Meier curve comparing patients without prior radiotherapy again demonstrated a statistically significant difference between the two groups, with a p of 0.0021 (Figure 3).

**Figure 3.**
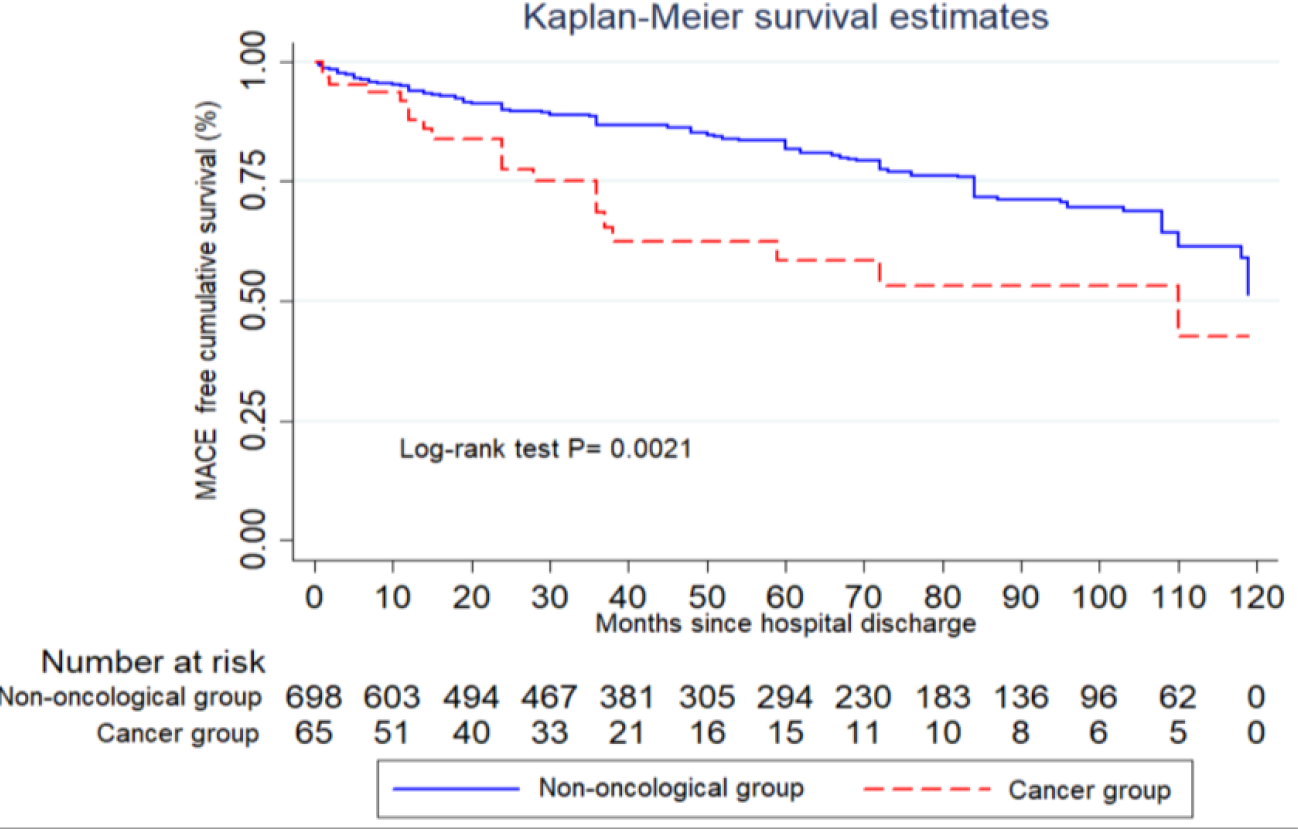
Kaplan-Meier Curve for Time to MACE comparing oncologic patients without prior Radiotherapy and non-oncologic patients.

The subgroup of patients without active chemotherapy consisted of 71 patients. The total number of MACE in this subgroup analysis was 165, with 147 events in G1 and 18 events in G2. The Kaplan-Meier curve comparing both subgroups showed p=0.043 (Figure 4). Cox Hazard analysis showed a non-significant difference between the incidence of MACE between the two groups, with an HR 1.61 (95% CI 0.97-2.68, p=0.066), adjusted for the risk factors mentioned above (Table S5)

**Figure 4.**
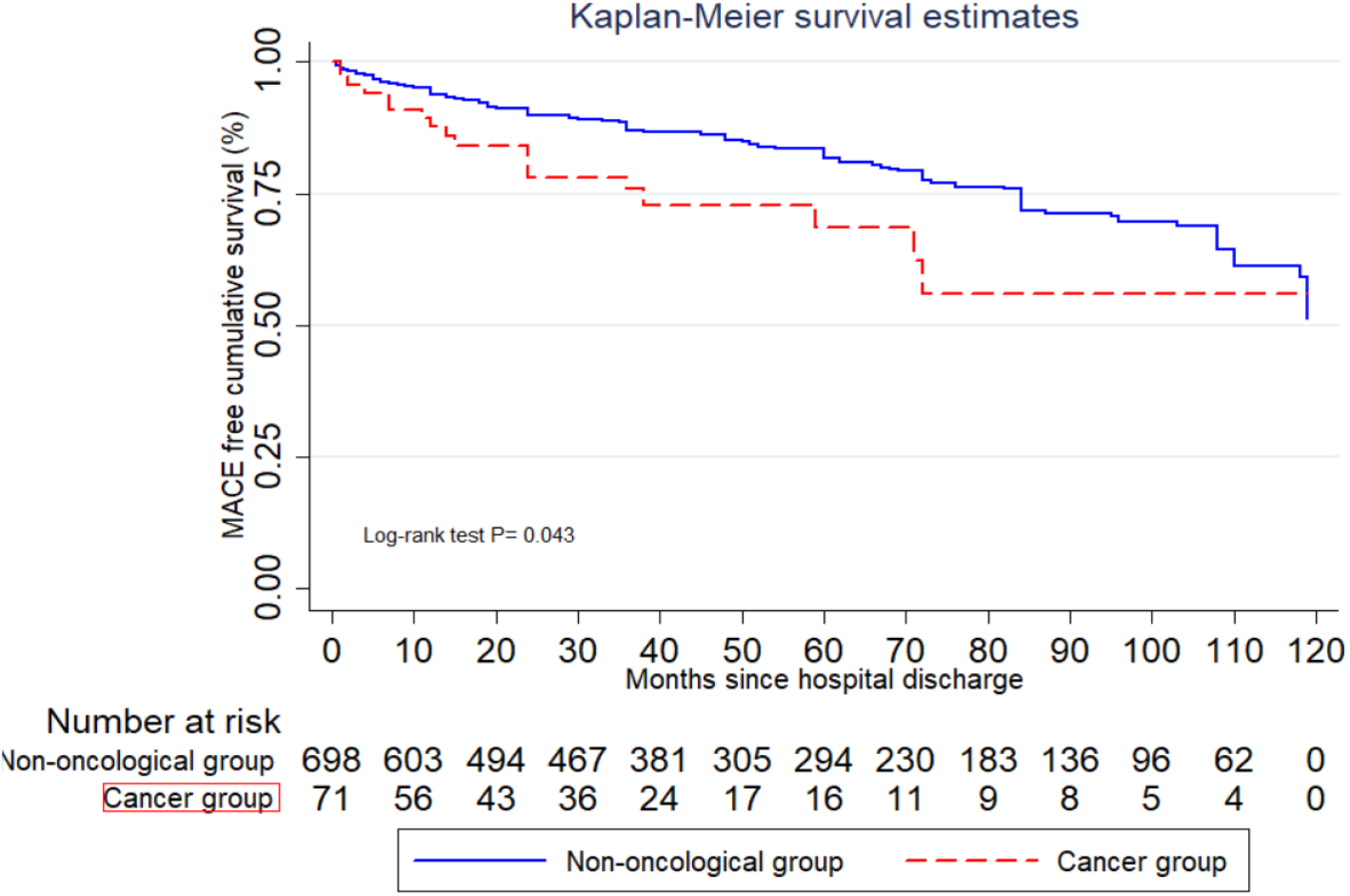
Kaplan-Meier Curve for Time to MACE comparing oncologic patients Without Active Chemotherapy and non-oncologic patients.

## DISCUSSION

The present work demonstrates that a history of cancer is an independent risk factor in the secondary prevention in our Latin American population, regardless of age at the time of ACS, gender, diabetes, high blood pressure, dyslipidemia, sedentary lifestyle, history of smoking and inherited family history. A first consideration to consider is that cancer patients have historically been poorly reflected in primary and secondary cardiovascular prevention studies due to systematic exclusion.

In our population, a history of cancer increased the risk of MACE by 84% during follow-up. The difference in rate of MACE in both groups during follow-up was mainly due to an increase in cardiovascular death and stroke; the other components of MACE were not statistically significant. Cardiovascular death could not be explained by differences in the therapeutic strategies carried out during hospitalization. The population with cancer received the same treatment as the general population, in accordance with current guidelines regarding the use of primary or rescue PCI as appropriate.

Left ventricular ejection fraction (LVEF) is an important factor for survival after a coronary event (22, 23), and at discharge, the LVEF was similar in both groups, indicating that an impaired LVEF at the time of discharge is unlikely to explain the association between cancer and MACE during follow-up in our cohort. Diabetes is also considered an independent risk factor for subsequent events as well (24, 25). In our cohort, diabetic cancer patients experienced 30% of events during follow-up, while diabetic patients without a history of cancer experienced 27% of events, with no statistically significant differences.

Recently, Tabata et al. (20) and Wright et al (26), assessed the incidence of major cardiovascular events in patients with a history of cancer in a secondary prevention context without considering the management strategies or care quality of secondary prevention. Our study is consistent with these findings despite population differences, but we explicitly explored that the association was not due to lower attainment of cardiovascular secondary prevention targets.

In recent years, an emphasis has been made on the impact of radiotherapy as a risk factor for coronary atherosclerosis (27-29). In our population, the subgroup analysis excluding the history of radiotherapy showed the same results as the general population of the cohort. The analysis of patients without active oncological treatment presented a similar trend, with the survival curves being different from each other, demonstrating a strong epidemiological implication without the exposure variable being an independent risk factor because it did not reach statistical significance.

Previous studies have suggested that there might be some direct endothelial compromise with selected cancer treatment in both pediatric and adult populations (9, 12-14). However, the rate of patients with active chemotherapy treatment was 20% (18 out of 89 patients), and in the subgroup analysis the relationship between cancer and worsened cardiovascular outcomes persisted. Therefore, in our results, this damage from the oncologic treatment performed would not have an impact on generating distortion of the events, giving greater value to the reported results. Patients with immunotherapies were underrepresented in our work, so the results were not due to the atherogenic potentiation of these drugs. In summary, our results enhance an association between oncological history and MACE beyond the cancer treatment strategy used. It would be necessary to elucidate the individual impact of each therapy, type of cancer and latencies to give a specific value to each situation and its future cardiovascular implication.

Currently, the inflammatory and multifactorial genesis of atheroma plaque is a well-established causal mechanism. Among the possible pathophysiological mechanisms that would explain the findings, we believe there would be a possible residual impact of endothelial inflammation linked to the oncological pathology and the treatments received. Cancer, with its pro-inflammatory mechanisms and the deleterious effects of its treatment, generates an increase in cytokines and pro-inflammatory factors that lead to an imbalance in endothelial function, such as a decrease in Nitric Oxide, an increase in Endothelin-1, Interleukin-6 and Tumor Growth Factors. (14, 30-33). Our study provides evidence that the association is likely real and not confounded by other determinants of subsequent events in the cardiovascular secondary prevention setting. That suggests a statistically significant and clinically relevant association between a history of cancer and the occurrence of cardiovascular events in secondary prevention. Our results hence suggest the need to explore if patients with a history of cancer should be considered for a stricter secondary prevention strategy, and further prospective studies are needed to evaluate the true role of cancer in this phase.

### Limitations

Our study was designed as a retrospective observational study and thus, data collection was limited to that available at the time of the study, providing an inferior level of evidence compared to a prospective study. Another limitation was that we adjusted for traditional cardiovascular risk factors such as hypertension, smoking, diabetes, etc., but we did not adjust for frailty, low BMI, anemia, autoimmune disorders, among other comorbidities that have been associated with worsening cardiovascular outcomes (34). Additionally, we did not seek for the presence of clonal hematopoiesis of indeterminate potential (CHIP) in our cohort due to insurance coverage limitations, despite being a condition associated with hematologic malignancy and increased cardiovascular risk. Finally, the goals for secondary prevention for our cohort, such as optimal LDL levels and statin doses, varied throughout our cohort given the rise of new guidelines with newer evidence.

## CONCLUSION

In our population, the incidence rate ratio of major cardiovascular events was higher in cancer patients. Oncological history behaved as an independent risk factor for major cardiovascular events, adjusted for classic risk factors. These results highlight future endpoints for cardiovascular prevention and public health in this population.

## Data Availability

All data was collected from electronic medical records from our university hospital, and was stored in a confidential manner. This data is kept safe by the corresponding author and can be make available for review upon request.

## ACKNOWLEDGMENTS

RM was the designer and coordinator of this study SDS, LS, RM, and PR all contributed to data collection. RM and SDS did the data analysis. RM, SDS, MR, MOF, AL collaborated equally to the writing of this manuscript.

## SUPPLEMENTARY FILES

Supplementary Tables 1-5

Supplementary Methods 1

